# Shrub and Forest Proximity and Cattle Farming Drive Tick (Acari: Ixodidae) Exposure Risk in the SFTS Endemic Region of Chongqing, China

**DOI:** 10.64898/2025.12.03.25341461

**Authors:** Ran Liu, Lijun Ran, Yan Zong, Liping Ran, Yucheng Qian, Taotian Tu, Yu Tan

**Author notes:** Corresponding Author: Taotian Tu (No. 187, Tongxing North Road, Beibei District, Chongqing 400707); Yu Tan (No. 23, Dudu Avenue, Wan’an Street, Shizhu Tujia Autonomous County, Chongqing 409100). These authors contributed equally to this work (Ran Liu, Lijun Ran).

## Abstract

Tick distribution in China has significantly expanded with urbanization and climate change. Chongqing faces a significant risk of severe fever with thrombocytopenia syndrome (SFTS), but targeted prevention efforts are challenging due to unclear exposure pathways. This study combined generalized linear model (GLM), Bayesian networks, and Bayesian multivariate GLM to assess environmental, agricultural, and socioeconomic drivers of tick exposure in southwestern China. Direct tick exposure risks primarily arose from proximity to shrublands, forests, and cattle farming. Pet ownership also increased risk, while proximity to croplands reduced exposure, likely due to the pesticide/herbicide use and tillage. Bayesian networks revealed that socioeconomic factors indirectly mediated risk. Higher education levels reduced cattle farming likelihood and increased income tier, lowering exposure by altering land-use proximity and agricultural activities. Key factors showing no significant association included demographics (age/gender), grassland proximity, and crop cultivation. Bayesian methods resolved collinearity and mediation effects in GLM, clarifying township-level tick exposure mechanisms in Southwest China and mapping driver networks. Findings demonstrate that tick exposure stems from complex interactions among environmental, agricultural, and socioeconomic factors. Prevention in mountainous southwest China should prioritize the shrubland/forest–cattle farming ecological interface. Future studies should integrate geospatial data for enhanced risk mapping.

## 1. Introduction

Ticks are notorious arthropod vectors responsible for spreading various detrimental diseases(Madison-Antenucci et al. 2020, Shah et al. 2023). With global warming, urbanization, and globalization, ticks have become a growing public health threat in China(Chen et al. 2014, Jia et al. 2022, Wu et al. 2024, Zhao et al. 2021, Zhou et al. 2014). Since 1980, ticks have significantly expanded their geographic distribution, spreading from northern regions to nearly all regions of the country and transmitting disease(Fang et al. 2015, Wu et al. 2024, Zhao et al. 2021). Among the pathogens transmitted by ticks, Lyme disease and tick-borne encephalitis have long been recognized as major concerns, particularly in North America and Europe(Madison-Antenucci et al. 2020, Shah et al. 2023). In contrast, East Asia, especially southern China, has witnessed the emergence of SFTSV, a novel bunyavirus with high case lethality rate around 7.8%(Cui et al. 2024, Seo et al. 2021, Sharma and Kamthania 2021, Wu et al. 2024). SFTSV is transmitted primarily through tick bites and secondary transmission via body fluids or aerosols(Kim and Park 2023). From 2010 to 2019, China has reported 13824 infections, and 713 deaths attributed to SFTS(Huang et al. 2021). Notably, in the last three years, confirmed SFTSV cases were identified in Chongqing, China(Hu et al. 2024). All 6 cases with 2 fatalities were reported in Xituo area, Shizhu County of Chongqing, highlighting a critical public health concern.

High-risk behaviors for tick exposure include prolonged outdoor activities in tick-infested shrub, forests, or grasslands(Fischhoff et al. 2019, Jore et al. 2020, SCHWARTZ and GOLDSTEIN 1990). A U.S. study by Wilson demonstrated that spending more than 7 hours per week in high-risk areas triples the likelihood of tick attachment(Wilson et al. 2023). Additionally, interactions with hunting dogs have been linked to elevated exposure risks(Sgroi et al. 2022, Toepp et al. 2018). While a study in northeastern China identified outdoor activities in forested or grass land as significant risk factors(Fang et al. 2024), these findings may not generalize to mountainous regions in Chongqing due to climatic, behavioral, and geographical differences. Crucially, mechanisms underlying tick exposure pathways and their socio-behavioral drivers remain understudied in southwest China, hindering targeted prevention strategies.

To address this gap, our study combined GLM, Bayesian networks, and Bayesian multivariate GLM to analyze questionnaire-derived data from residents in Chongqing SFTS endemic region. By examining sociodemographic, environmental, and behavioral variables, we aim to unravel the causal chains of tick exposure and propose evidence-based interventions tailored to Chongqing’s unique ecological landscape.

## 2. Materials and methods

Ethical approval was obtained from the Human Research Ethics Committee at the Chongqing Center for Diseases Prevention and Control (KY-2025-030-1).

### 2.1 Study population

This study recruited residents from six townships in the Xituo area of Shizhu County, Chongqing. Five of these townships (Xituo, Wanchao, Lichang, Wangchang, and Yanxi) are within 2 kilometers of the Yangtze River. Yuchi Town is located 15 kilometers northwest of the Yangtze River and is adjacent to Qianye Grassland (a protected grassland area and scenic spot).

### 2.2 Questionnaire Design and data collection

A branching-logic questionnaire was developed in Mandarin Chinese, structured into four domains, demographic characteristics, environmental factors, biting incident profile, health belief model assessment. Given the study’s focus on environmental exposure determinants, core analytical variables included:

1. Demographic information: age (free entry); gender; education level; living town; living in rural area or urban area.
2. Exposure factors: whether feeding livestock, if yes, include which livestock (swine, cattle, duck/goose, goat/sheep, chicken); whether feeding pet, if yes, include which pet (feline, canine); whether found ticks in their living environment in last 3 years; which places their living environment includes in 500 diameters (shrub, forest, grassland, cropland, ranch).

From March 2025 to April 2025, as part of the disease prevention efforts at the Shizhu County Center for Disease Control and Prevention, online questionnaires were administered to residents by primary health care providers through home visits, telephone interviews, or community gatherings. Participants had the option of completing it themselves or with assistance (e.g., for elderly or illiterate participants).

### 2.3 Data analysis

All analysis were conducted in R (version 4.4.3) and RStudio (version 2024.12.1). Questionnaires data cleaning and transformation were executed, retaining only questionnaires where participants answered “Yes” to the question “Are you able to identify ticks?” for further analysis. Participants’ demographic and exposure-related characteristics were summarized using descriptive statistics, with categorical variables reported as percentages. Age was categorized into three groups (≤35 years, 35–65 years, >65 years). Group differences between non-exposure and exposure participants were assessed using Pearson’s chi-squared test for categorical variables. To identify direct predictors of tick exposure, a binary logistic regression model was constructed. Covariates included demographic variables (age, gender, education, income, living place (town)), environmental factors (proximity of shrubs, grasslands, ranch, forest and livestock/pet ownership). And collinearity was checked using variance inflation factors. To mitigate overfitting risks from an overly complex model, only variables hypothesized a priori to directly influence environmental contact (e.g., livestock ownership, shrub proximity) were retained, specific livestock types (e.g., cattle, swine) and pet types (e.g., canine) were not considered.

To disentangle the interdependencies between variables and the pathways of exposure, Bayesian network analysis was performed using the bnlearn package (v5.0.2)(Scutari 2010). The predictive strength of the variables was assessed using the bootstrap method (1,000 iterations), and the directed acyclic graph (DAG) was visualized using the Rgraphviz package (v2.50.0)(Scutari 2010). The initial network incorporated all variables, but only those directly or indirectly linked to exposure status or demonstrating a significant effect (*P*<0.05) on exposure status in prior generalized model analysis were retained in the final network. Network structure was constrained by two principles grounded in causal plausibility: (1) living place (town) was fixed as a parent node and exposure status was fixed as a child node to reflect causal precedence; (2) Direct arc was blocked from place (town) to exposure status to represent the outcome of interest. The threshold of 0.5 was also set for both Bayesian networks. Subsequently, the posterior distributions of key nodes in the final network were estimated using a Bayesian multivariate GLM by brms package (v2.22.0)(Bürkner 2017), enabling probabilistic inference on exposure pathways. This model used 4 chains, each with 2,000 iterations (1,000 warm-up), no thinning, and yielded 4,000 post-warmup draws.

## 3. Result

### 3.1 Participant Characteristics

A total of 902 questionnaires were collected. Following quality checks and data cleaning, 126 responses (58 incomplete or duplicate, 67 unable to identify ticks) were excluded, resulting in 777 valid questionnaires retained for analysis (Figure 1).

**Figure 1.**
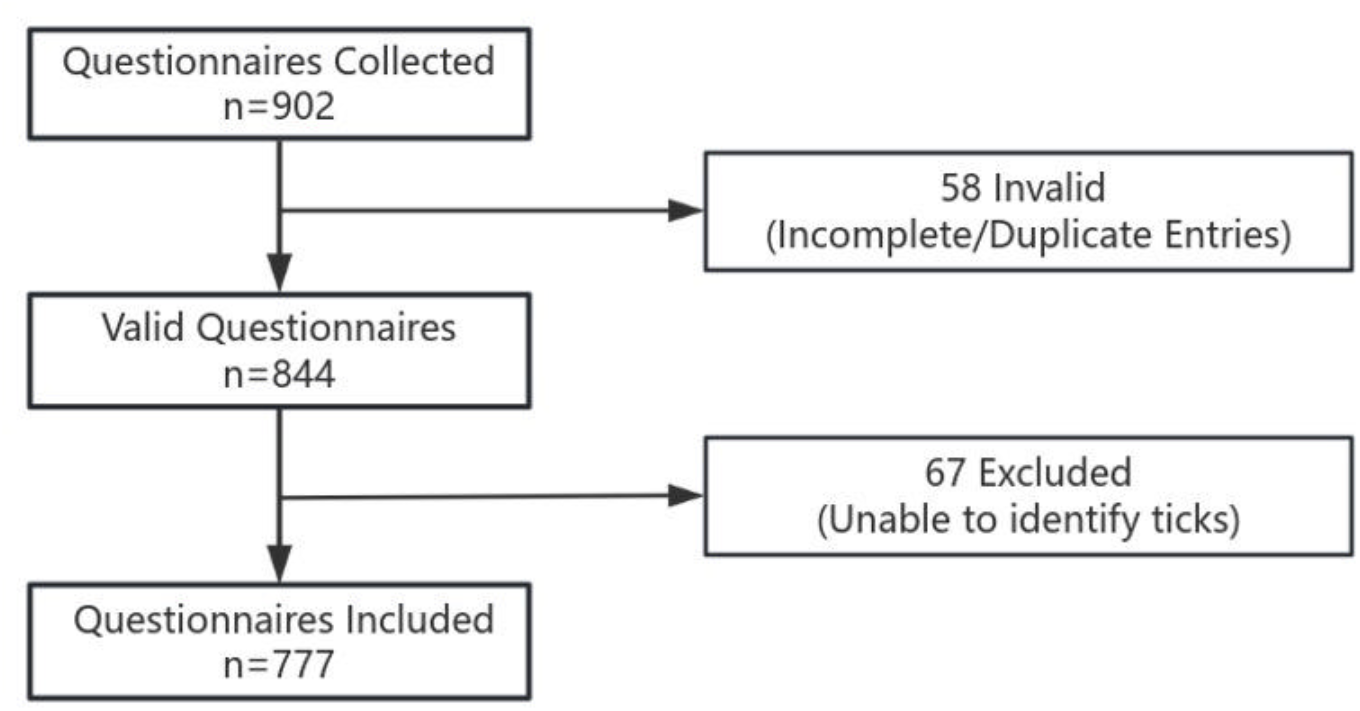
Flow chart of questionnaire collection

The demographic characteristics and exposure factors of the exposed and non-exposed participants in this study were shown in Table 1. In total, there were 370 (47.6%) in the exposed group and 407 (52.4%) in the non-exposed group. Chi-square tests revealed significant age distribution differences (χ^2^=14.315, df=2, P<0.001), showing lower exposure rates among those ≤35 years and higher rates in the 35-65. Education level differed significantly between groups (χ^2^=22.443, df=3, P<0.001), with 74.9% of exposed participants having primary education versus 60.7% in non-exposed. Income disparities were evident with 78.6% of exposed households earning <3000 CNY monthly compared to 61.9% non-exposed (χ^2^=26.052, df=2, P<0.001). Geographical analysis showed substantial residential variation (χ^2^=64.542, df=5, P<0.001), particularly in Wanchao and Yanxi. Tick exposure was significantly (P<0.001)) associated with cattle ownership and poultry ownership. Pet ownership patterns demonstrated elevated exposure risks for canine and feline. Environmental factors showed increased exposure with forest proximity (χ^2^=34.848, df=1, P<0.001), shrubland proximity (χ^2^=90.963, df=1, P<0.001) and grassland proximity (χ^2^=11.363, df=1, P<0.001), while cropland proximity exhibited inverse association (χ^2^=16.825, df=1, P<0.001).

**Table 1.** Descriptive analysis of questionnaires.

| Variable | Exposure | | Total<br>n(%) | $\chi^2$ | df | P value |
| --- | --- | --- | --- | --- | --- | --- |
|  | No<br>n(%) | Yes<br>n(%) |  |  |  |  |
| Age |  |  |  |  |  |  |
| $\leq 35$ | 27 (6.6%) | 8 (2.2%) | 35 (4.5%) | | | |
| 35-65 | 181 (44.5%) | 203 (54.9%) | 384 (49.4%) | 14.315 | 2 | $<0.001$ |
| $>65$ | 199 (48.9%) | 159 (43%) | 358 (46.1%) | | | |
| Gender |  |  |  |  |  |  |
| Male | 323 (79.4%) | 296 (80%) | 619 (79.8%) | 0.017 | 1 | .895 |
| Female | 84 (20.6%) | 74 (20%) | 158 (20.3%) |  |  |  |
| Education |  |  |  |  |  |  |
| Primary | 247 (60.7%) | 277 (74.9%) | 524 (67.4%) | 22.443 | 3 | <0.001 |
| Junior | 107 (26.3%) | 64 (17.3%) | 171 (22.1%) |  |  |  |
| High | 21 (5.2%) | 19 (5.1%) | 40 (5.2%) |  |  |  |
| College | 32 (7.9%) | 10 (2.7%) | 42 (5.4%) |  |  |  |
| Monthly family income (CNY) |  |  |  |  |  |  |
| <3,000 | 252 (61.9%) | 291 (78.6%) | 543 (69.9%) | 26.052 | 2 | <0.001 |
| 3,000-6,000 | 130 (31.9%) | 64 (17.3%) | 194 (25.0%) |  |  |  |
| >6,000 | 25 (6.1%) | 15 (4.1%) | 40 (5.2%) |  |  |  |
| Living place (town) |  |  |  |  |  |  |
| Lichang | 50 (12.3%) | 48 (13%) | 98 (12.6%) | 64.542 | 5 | <0.001 |
| Wanchao | 26 (6.4%) | 72 (19.5%) | 98 (12.6%) |  |  |  |
| Wangchang | 68 (16.7%) | 64 (17.3%) | 132 (17.0%) |  |  |  |
| Xituo | 46 (11.3%) | 59 (15.9%) | 105 (13.5%) |  |  |  |
| Yanxi | 166 (40.8%) | 67 (18.1%) | 233 (30.0%) |  |  |  |
| Yuchi | 51 (12.5%) | 60 (16.2%) | 111 (14.3%) |  |  |  |
| Crop cultivation |  |  |  |  |  |  |
| No | 71 (17.4%) | 52 (14.1%) | 123 (15.8%) | 1.4275 | 1 | .232 |
| Yes | 336 (82.6%) | 318 (85.9%) | 654 (84.2%) |  |  |  |
| Livestock husbandry |  |  |  |  |  |  |
| No | 128 (31.4%) | 59 (15.9%) | 187 (24.1%) | 29.941 | 1 | <0.001 |
| Yes | 279 (68.6%) | 311 (84.1%) | 590 (75.9%) |  |  |  |
| Cattle ownership |  |  |  |  |  |  |
| No | 194 (47.7%) | 117 (31.6%) | 311 (40.0%) | 24.65 | 1 | <0.001 |
| Yes | 213 (52.3%) | 253 (68.4%) | 466 (60.0%) |  |  |  |
| Goat/Sheep ownership |  |  |  |  |  |  |
| No | 350 (86%) | 321 (86.8%) | 671 (86.4%) | 0.042 | 1 | .838 |
| Yes | 57 (14%) | 49 (13.2%) | 106 (13.6%) |  |  |  |
| Swine ownership |  |  |  |  |  |  |
| No | 307 (75.4%) | 235 (63.5%) | 542 (69.8%) | 12.486 | 1 | <0.001 |
| Yes | 100 (24.6%) | 135 (36.5%) | 235 (30.2%) |  |  |  |
| Chicken ownership |  |  |  |  |  |  |
| No | 313 (76.9%) | 208 (56.2%) | 521 (67.1%) | 36.616 | 1 | <0.001 |
| Yes | 94 (23.1%) | 162 (43.8%) | 256 (33.0%) |  |  |  |
| Duck/goose ownership |  |  |  |  |  |  |
| No | 350 (86%) | 247 (66.8%) | 597 (76.8%) | 39.226 | 1 | <0.001 |
| Yes | 57 (14%) | 123 (33.2%) | 180 (23.2%) |  |  |  |
| Pet ownership |  |  |  |  |  |  |
| No | 301 (74%) | 180 (48.6%) | 481 (61.9%) | 51.566 | 1 | <0.001 |
| Yes | 106 (26%) | 190 (51.4%) | 296 (38.1%) |  |  |  |
| Canine ownership |  |  |  |  |  |  |
| No | 311 (76.4%) | 201 (54.3%) | 512 (65.9%) | 41.099 | 1 | <0.001 |
| Yes | 96 (23.6%) | 169 (45.7%) | 265 (34.1%) |  |  |  |
| Feline ownership |  |  |  |  |  |  |
| No | 369 (90.7%) | 294 (79.5%) | 663 (85.3%) | 18.548 | 1 | <0.001 |
| Yes | 38 (9.3%) | 76 (20.5%) | 114 (14.7%) |  |  |  |
| Urban/rural |  |  |  |  |  |  |
| Urban | 22 (5.4%) | 11 (3%) | 33 (4.2%) | 2.253 | 1 | .133 |
| Rural | 385 (94.6%) | 359 (97%) | 744 (95.8%) |  |  |  |
| Environment |  |  |  |  |  |  |
| Forest proximity |  |  |  |  |  |  |
| No | 263 (64.6%) | 160 (43.2%) | 423(54.44%) | 34.848 | 1 | <0.001 |
| Yes | 144 (35.4%) | 210 (56.8%) | 354(45.56%) |  |  |  |
| Cropland proximity |  |  |  |  |  |  |
| No | 56 (13.8%) | 95 (25.7%) | 151(19.43%) | 16.825 | 1 | <0.001 |
| Yes | 351 (86.2%) | 275 (74.3%) | 626(80.57%) |  |  |  |
| Shrub proximity |  |  |  |  |  |  |
| No | 326 (80.1%) | 174 (47%) | 500(64.35%) | 90.963 | 1 | <0.001 |
| Yes | 81 (19.9%) | 196 (53%) | 277(35.65%) |  |  |  |
| Grassland proximity |  |  |  |  |  |  |
| No | 213 (52.3%) | 148 (40%) | 361(46.46%) | 11.363 | 1 | <0.001 |
| Yes | 194 (47.7%) | 222 (60%) | 416(53.54%) |  |  |  |
| Park proximity |  |  |  |  |  |  |
| No | 395 (97.1%) | 358 (96.8%) | 753(96.91%) | <0.001 | 1 | .976 |
| Yes | 12 (2.9%) | 12 (3.2%) | 24(3.09%) |  |  |  |
| Ranch proximity |  |  |  |  |  |  |
| No | 350 (86.0%) | 274 (74.1%) | 624(80.31%) | 16.728 | 1 | <0.001 |
| Yes | 57 (14.0%) | 96 (25.9%) | 153(19.69%) |  |  |  |

**Table 2.**
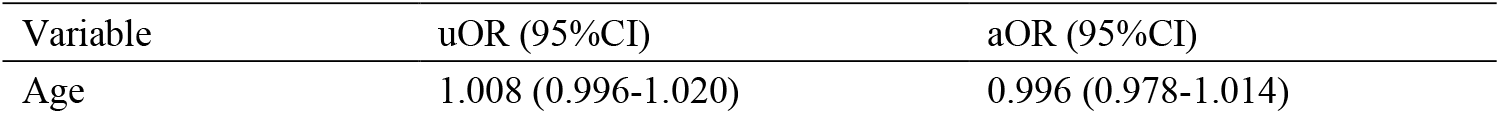
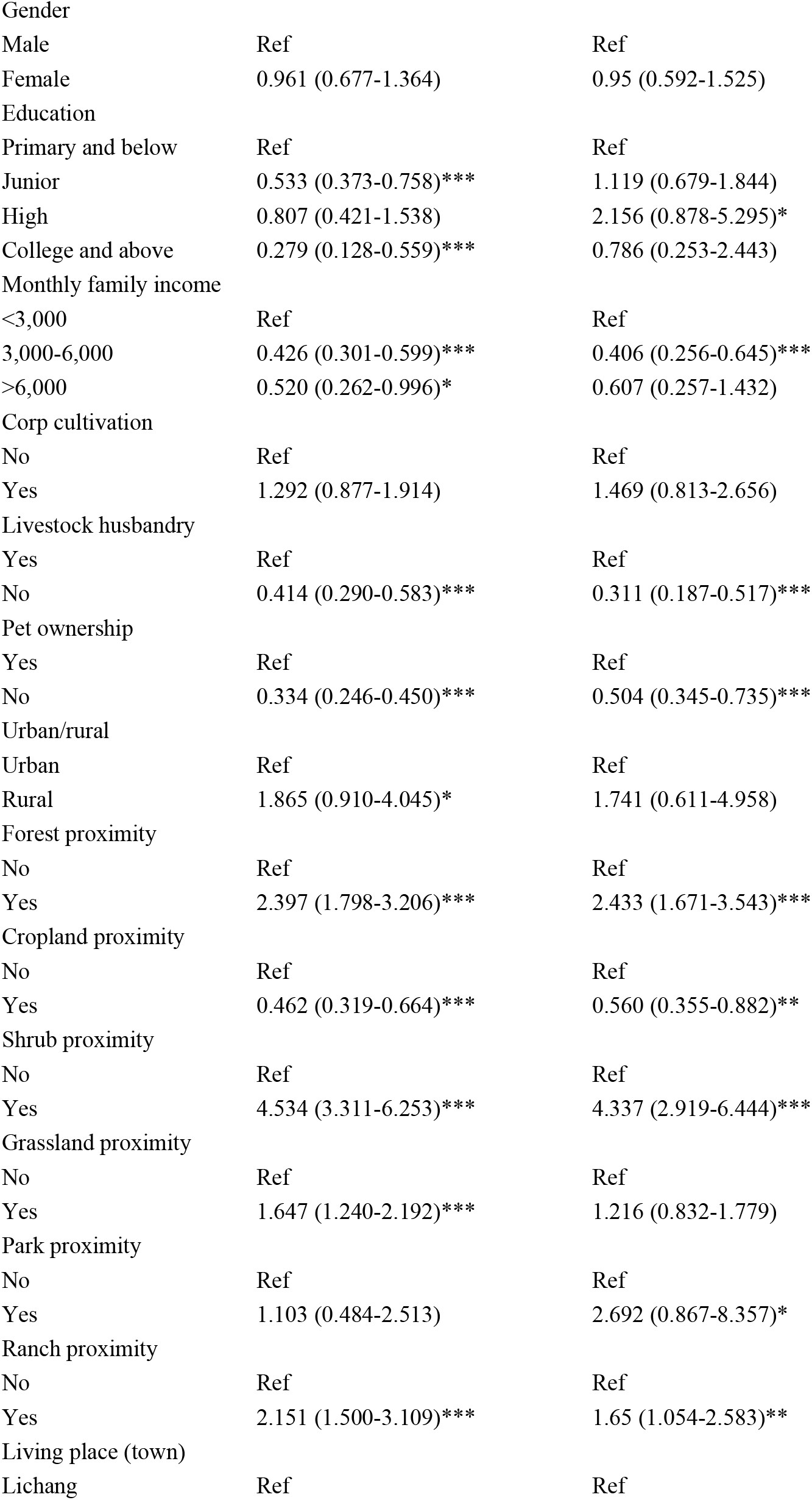
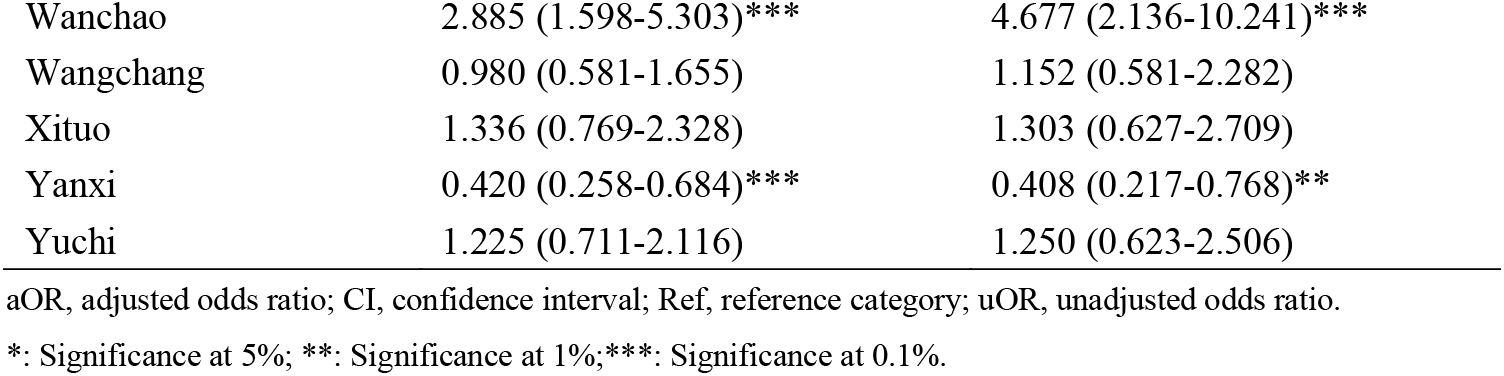
Result of GLM analysis.

### 3.2 GLM risk factor analysis

Significant associations between tick exposure and risk factors were identified through unadjusted (uOR) and adjusted odds ratios (aOR) (Table 1). Demographic analysis revealed no age-related risk or gender disparity. Middle-income households (3,000–6,000 CNY/month) exhibited reduced risk (aOR=0.406, P<0.001) compared to lower-income groups. Not having livestock conferred protection (aOR = 0.311, P<0.001), while not having pets was also associated with a reduced risk (aOR = 0.504, P<0.001). Environmental factors showed shrubland proximity as the strongest risk factor (aOR=4.337, P<0.001), contrasting with cropland’s protective effect (aOR=0.560, p value <0.01). Geospatial heterogeneity was marked, with Wanchao residents facing 4.7-fold elevated risk (aOR=4.677, P<0.001) versus Yanxi’s reduced risk (aOR=0.408, p value<0.01). Model diagnostics indicated acceptable multicollinearity (VIF<2.82). The significance shifts between uOR and aOR suggest potential confounding by education and grassland exposure, underscoring the necessity of adjustment in causal inference.

### 3.3 Bayesian network structure

The initial Bayesian network (Figure 2) was constructed using a model averaging approach. Key dependencies revealed socioeconomic and agricultural interactions. Education directly influenced crop cultivation (strength=0.650) and cattle ownership (strength=0.813), while town served as a hub for environmental variables (shrub, grassland) and livestock (goat). Cattle ownership exhibited relationships with multiple variables and served as a central node in exposure pathways. Environmental factors like shrub strongly predicted exposure (strength = 0.999), and crop cultivation drove downstream outcomes (income, rural/urban). Notably, feedback loops emerged in forest-grassland and swine-chicken-feline interactions, highlighting dynamic interdependencies. While the initial network omitted the arc from chicken to swine due to cyclic dependencies in the DAG, this oversight is unlikely to significantly impact the study, as the excluded edge had negligible effects on the overall pathway.

**Figure 2.**
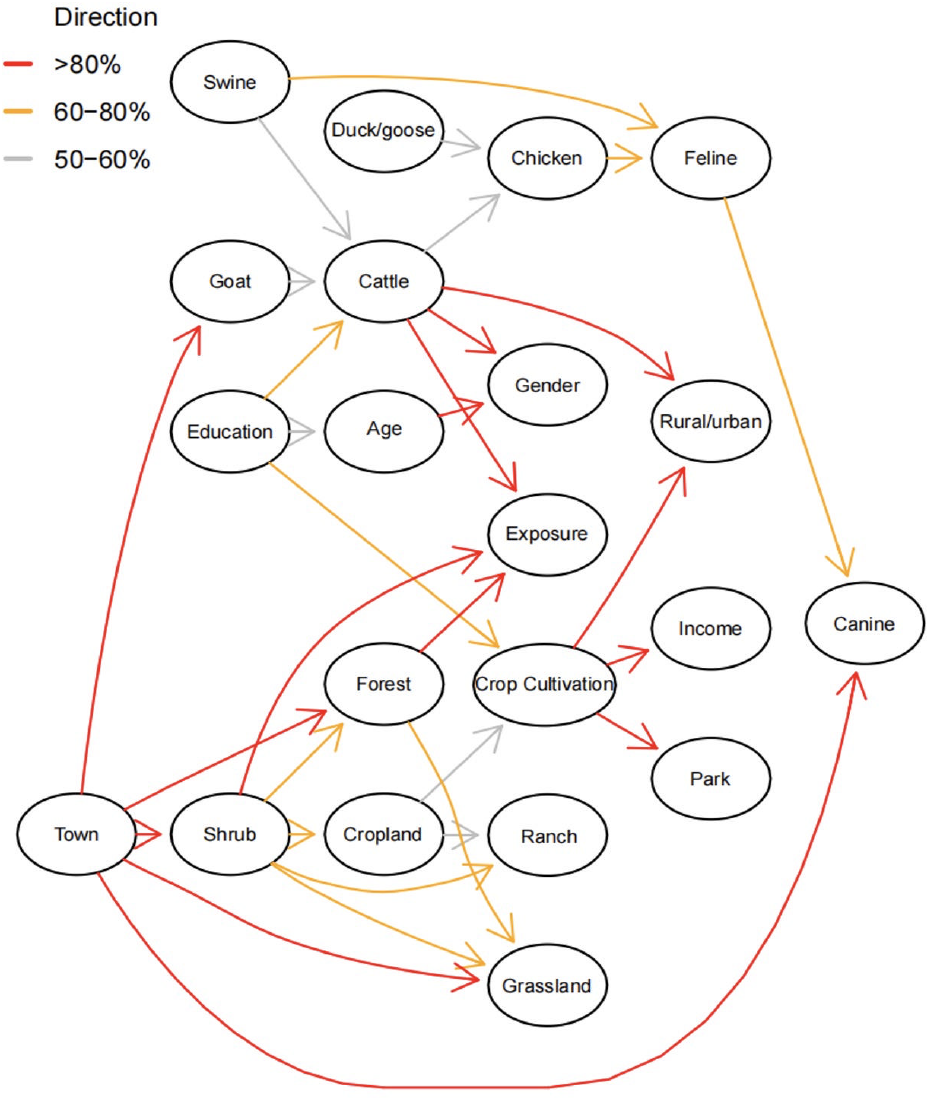
Initial Bayesian network

Figure 3 depicts the final refined Bayesian network. The relationship between swine ownership and pet ownership shifted from swine ownership to feline ownership to swine ownership to canine ownership in the final network. Notably, town’s governance no longer directly drove canine ownership but instead acted through feline ownership (strength=0.999), which interacted with canine ownership (strength=1.000). The remaining framework for environmental factors remained unchanged, with forest proximity and shrub proximity collectively driving exposure (strengths=0.695 and 1.000).

**Figure 3.**
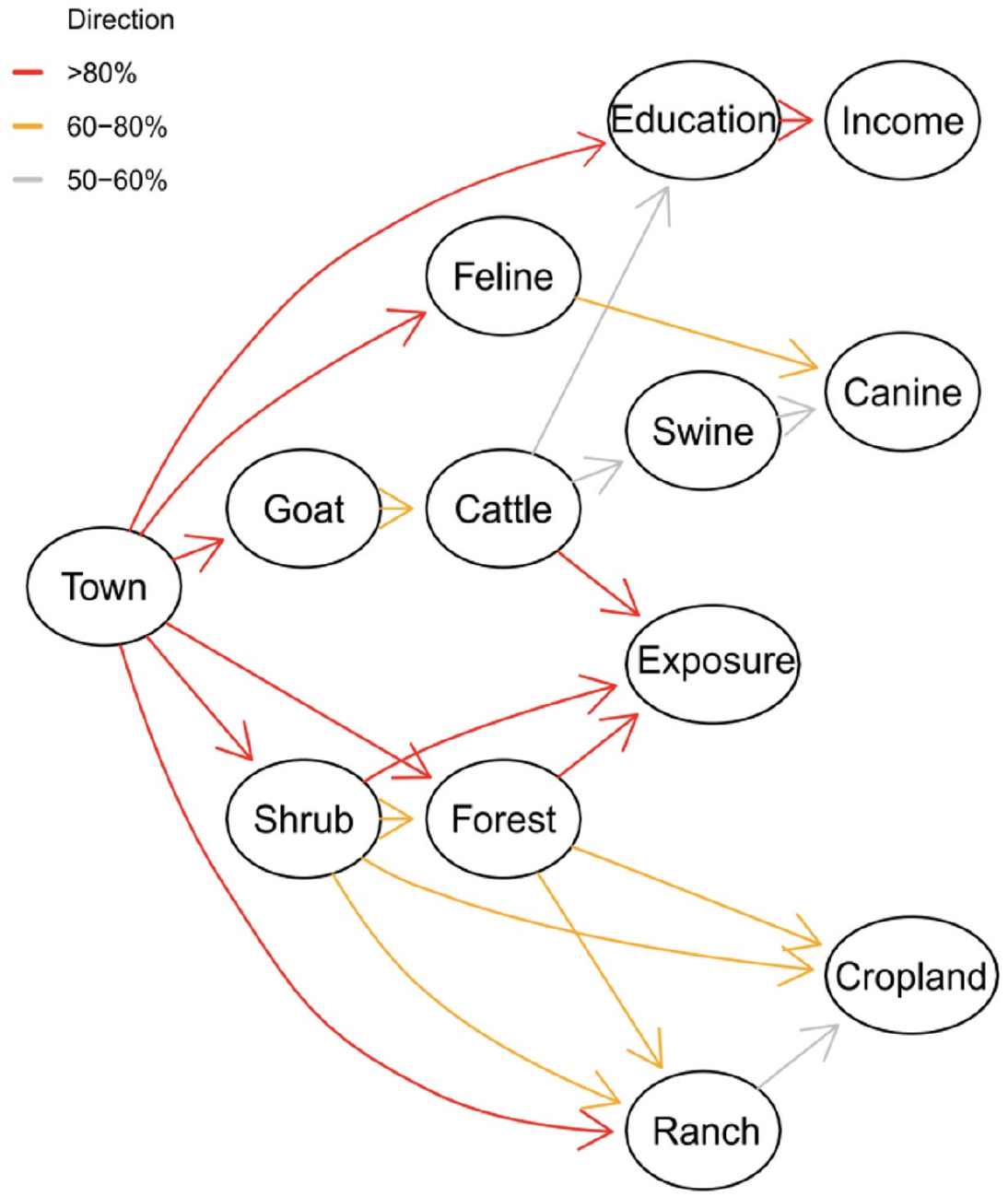
Final Bayesian network

### 3.4 Bayesian multivariate GLM model

Key nodes from the final Bayesian network were integrated into a Bayesian multivariate GLM with fixed effects (Figure 4). Education exhibited town-level heterogeneity. Junior school attainment was inversely associated with residence in Wanchao (Bayesian OR=0.013, 95%CI=0.002–0.053) and Xituo (Bayesian OR=0.154, 95%CI=0.076–0.304), while college-level education showed elevated odds in Wangchang (Bayesian OR=5.266, 95%CI=1.15-35.25). Cattle ownership reduced the likelihood of higher education (college: Bayesian OR=0.027, 95%CI=0.007–0.082). Income disparities were strongly mediated by education, college attainment amplified odds of earning >6,000 CNY (Bayesian OR=16.78, 95%CI=5.81–46.67) compared to junior school (Bayesian OR=2.49, 95%CI=1.11–5.26).

**Figure 4.**
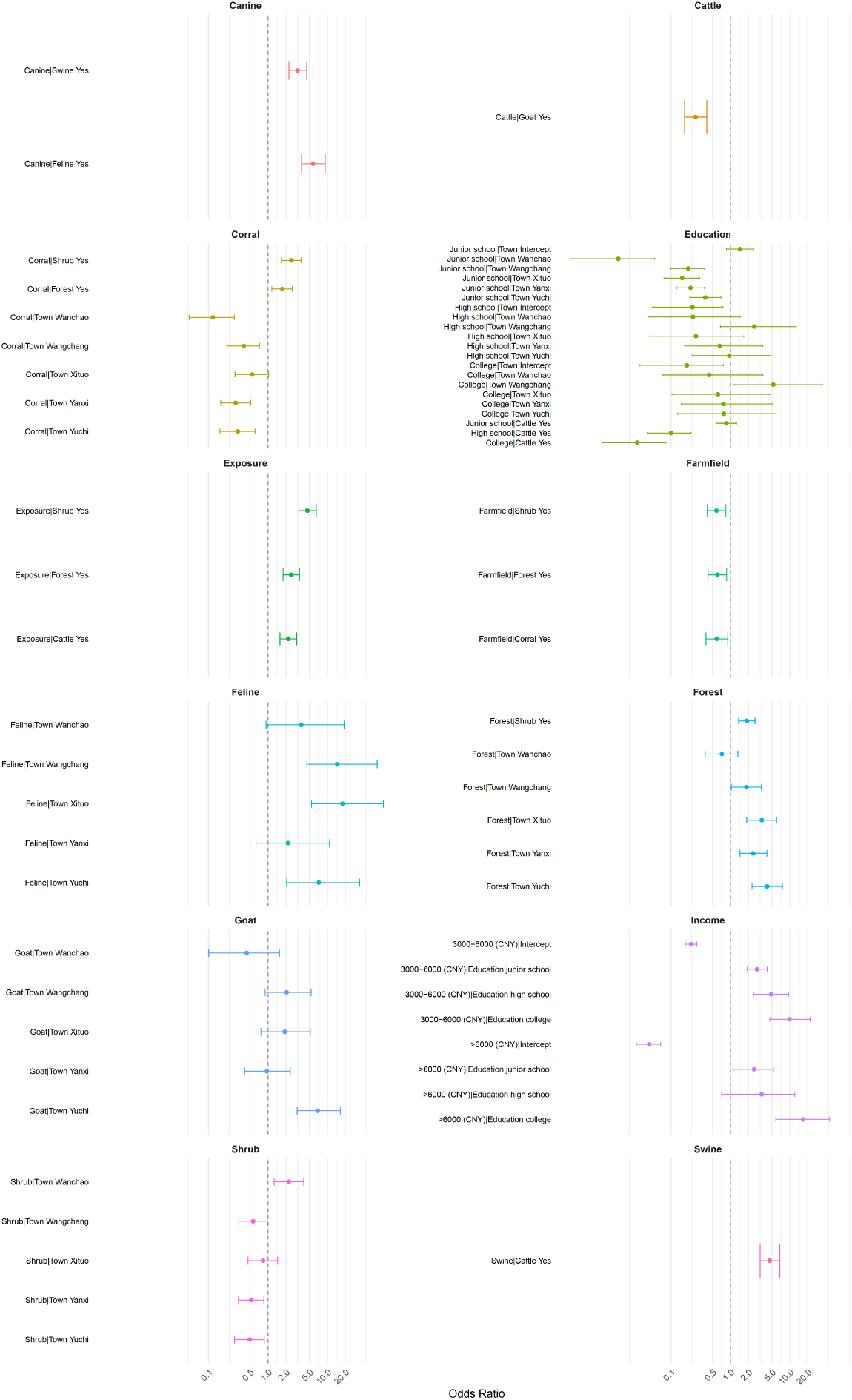
Result of Bayesian Multivariate GLM analysis

Exposure was strongly associated with shrub proximity (Bayesian OR=4.601, 95%CI=3.314–6.476), forest proximity (Bayesian OR=2.448, 95%CI=1.784–3.386), and cattle ownership (Bayesian OR=2.195, 95%CI=1.589–3.052), with shrub proximity exhibiting the highest posterior odds ratio. Livestock patterns revealed spatial clustering, Yuchi had heightened goat ownership (Bayesian OR=6.84, 95%CI=3.10–16.47), while feline ownership peaked in Xituo (Bayesian OR=17.92, 95%CI=5.38–87.28) and Wangchang (Bayesian OR=14.60, 95%CI=4.52–68.70). Cattle ownership predicted swine ownership (Bayesian OR=4.58, 95%CI=3.16–6.76) and canine ownership co-occurrence (Bayesian OR=3.15, 95%CI=2.24–4.47). Shrub proximity distribution varied markedly by town. Wanchao had elevated shrub proximity coverage (Bayesian OR=2.239, 95%CI=1.256– 3.982) but suppressed or unchanged elsewhere. Forest proximity displayed synergistic relationships with shrub proximity (Bayesian OR=1.886, 95%CI=1.367–2.592) and geographic clustering, peaking in Xituo (OR=3.371, 95%CI=1.878–5.999) and Yuchi (Bayesian OR=4.143, 95%CI=2.302–7.480). Ranch proximity was linked to shrub proximity (Bayesian OR=2.474, 95%CI=1.683–3.626) and forest proximity (Bayesian OR=1.739, 95%CI=1.156–2.565) but sharply reduced in Wanchao (Bayesian OR=0.118, 95%CI=0.049–0.271) and Yanxi (Bayesian OR=0.286, 95%CI=0.160–0.512). Cropland proximity declined with shrub proximity (OR=0.583, 95%CI=0.406–0.835), forest proximity (Bayesian OR=0.604, 95%CI=0.417–0.863), and ranch proximity (Bayesian OR=0.591, 95%CI=0.388–0.902), indicating land-use patterns. Model exhibited robust convergence and reliability (Rhat<1.01, with Bulk/Tail ESS>1000).

## 4. Discussion

Combined analysis revealed that tick exposure risk is shaped by joint effects of environmental, agricultural, and socioeconomic factors. Both methods consistently identified direct predictors, proximity to shrubland, proximity to forest, and cattle husbandry. These findings further linked livestock farming and vegetation interactions to tick exposure(Hurtado and Giraldo-Ríos 2018, Mathisson et al. 2021, Rehman et al. 2017). Socioeconomic disparities exhibited divergent patterns across methods. Education showed null effects in GLM but structured risk hierarchically in Bayesian networks and Bayesian multivariate GLM, where higher education reduced cattle ownership (Bayesian OR 95%CI=0.027–0.10), elevated income tiers (Bayesian OR 95%CI=9.85– 16.78 for college to >6,000 CNY), and linked to risk-enabling land use. Bayesian pathways anchored education’s indirect role, revealing how it stratifies income and livestock access, which was masked in GLM. These findings align with broader evidence linking higher education and income to reduced tick exposure risk in underdeveloped area settings(Godfrey and Randolph 2011, íumilo et al. 2008). However, this contrasts with findings in metropolitan areas, where higher education may lead to increased outdoor recreational activities and, consequently, tick exposure.(Dernat and Johany 2019, Godfrey and Randolph 2011, Jore et al. 2020).

The other demographic covariates (age, gender, rural/urban) showed no significant effect on tick exposure. While our GLM results indicated no significant difference in tick exposure risk between rural and urban living places, the questionnaires were collected in towns within the Xituo area, where urbanization and vegetation land use are limited. This context may reflect conditions in many underdeveloped towns, but it might not adequately differentiate between metropolitan and rural settings, particularly as other studies has identified significant disparities of exposure risk in between(Bayles et al. 2013, Fang et al. 2024, Gould et al. 2024, Natsuaki 2021).

Also, it was found that while cropland proximity was suppressed by other vegetation variables (forest and shrub) and ranch proximity, the exposure risk of crop cultivation showed no significant change in our GLM (aOR=1.469, *P*=0.203). Perhaps, even with the potential use of pesticides and clearing of other vegetation near croplands, farmers may still need to traverse high-risk areas to reach their cropland. Additionally, our Bayesian network suggests farmers involved in crop cultivation may have lower education levels, mediated by risk factors like cattle ownership. Ranch proximity in the GLM had a significant effect on exposure risk, which may be mediated by high-risk vegetation factors (forest proximity and shrub proximity) in the Bayesian network. This aligns with routine tick surveillance data showing frequent tick proximity near ranch with infested livestock, as well as findings from a study in Malaysia(Ghane Kisomi et al. 2016). Interestingly, grassland, which is commonly assumed to be a high-risk factor for tick exposure, including in local surveillance, showed no significant effect in the GLM (aOR=1.216, *P*=0.312). The surveyed residents might rarely visit or spend limited time in these areas, and grasslands comprise a smaller proportion of the landscape compared to other vegetation types. Additionally, since grazing activities for cattle and goats in the study region occur primarily in forest and shrub dominated mountainous areas, people may have fewer opportunities for grassland exposure. Some research has also indicated that *Haemaphysalis longicornis*, the major tick species captured in our routine surveillance, infests primarily shrubs and rarely grasslands in north China(Zheng et al. 2012). However, this remains debated and may be attributed to factors like seasonality or other variables(Mathisson et al. 2021).

Goat husbandry emerged as a mediating node between town and cattle ownership, with goats exerting a downward effect on cattle (Bayesian OR=0.259, 95%CI=0.169–0.4). While cattle ownership showed a positive association with swine ownership in the Bayesian network, the directionality strength was modest (0.545), indicating mutual interference. The preferential ownership of grazing livestock may explain the inhibitory effect of goats on cattle farming, as smallholders (the primary livestock actors in our study) may specialize in raising only one grazing animal (cattle/goats), while pig farming occurs in closed corrals. The effect of pet ownership on exposure risk in the GLM did not align with the Bayesian network results, potentially due to indirect mediation effects from cattle/swine ownership or unmeasured factors like dog-walking and hunting activities (though not analyzed in our study)(Aenishaenslin et al. 2017, Dernat and Johany 2019), contrast with many that found no significant difference of exposure in pet ownership in general public analysis(Jore et al. 2020, Wilson et al. 2023). From the Bayesian method, it can be inferred that livestock-raising households may tend to keep dogs, thus creating an indirect risk pathway. And cattle can introduce ticks into domesticated environments through the egg-laying activities of ticks infesting them, while cats or dogs, acting as secondary hosts, may further facilitate the spread of ticks into human habitations(Zhang et al. 2014).

In conclusion, combination of both methods exposed mechanistic trade-offs. Critical consensus emerged for environmental drivers (shrub proximity/forest proximity) and cattle farming, while discrepancies in income and education underscored context-dependent interpretation. Public health intervention should thus be focused on theses core factors. However, our findings are constrained by self-reported exposure data, which may introduce recall bias, and the absence of geospatial vegetation metrics or livestock population densities. The sample was limited to less urbanized areas, which hinders the generalization of conclusions to settings with clear urban-rural divides. Future studies should integrate geospatial data and longitudinal designs to disentangle the interplay between land use, livestock practices, and zoonotic risk in transitioning economies.

## Data Availability

All data produced in the present study are available upon reasonable request to the authors.

## Acknowledgments

We thank the primary healthcare providers and local officer from towns of Xituo, Yuchi, Wanchao, Yanxi, Lichang and Wangchang for help the survey complement. And this study was supported by the Science-Health Joint Medical Scientific Research Project of Chongqing (Grant Number: 2026MSXM109).

## Author Contributions

Ran Liu: Conceptualization, Data curation, Formal analysis, Software, Methodology, Validation, Visualization, Writing – original draft, Writing – review & editing; Lijun Ran: Conceptualization, Investigation, Project administration, Resources, Writing – review & editing; Yan Zong: Investigation, Project Administration, Data Curation; Liping Ran: Investigation, Resources; Yucheng Qian: Resources; Taotian Tu: Funding acquisition, Supervision; Yu Tan: Funding acquisition, Supervision.

